# Comparison of Bayesian Networks, G-estimation and linear models to estimate causal treatment effects in aggregated N-of-1 trials with carry-over effects

**DOI:** 10.1101/2022.07.21.22277832

**Authors:** Thomas Gärtner, Juliana Schneider, Bert Arnrich, Stefan Konigorski

## Abstract

The aggregation of a series of N-of-1 trials presents an innovative and efficient study design, as an alternative to traditional randomized clinical trials. Challenges for the statistical analysis arise when there is carry-over or complex dependencies of the treatment effect of interest.

In this study, we evaluate and compare methods for the analysis of aggregated N-of-1 trials in different scenarios with carry-over and complex dependencies of treatment effects on covariates. For this, we simulate data of a series of N-of-1 trials for Chronic Nonspecific Low Back Pain based on assumed causal relationships parameterized by directed acyclic graphs. In addition to existing statistical methods such as regression models, Bayesian Networks, and G-estimation, we introduce a carry-over adjusted parametric model (COAPM).

The results show that all evaluated existing models have a good performance when there is no carry-over and no treatment dependence. When there is carry-over, COAPM yields unbiased and more efficient estimates while all other methods show some bias in the estimation. When there is known treatment dependence, all approaches that are capable to model it yield unbiased estimates. Finally, the efficiency of all methods decreases slightly when there are missing values, and the bias in the estimates can also increase.

This study presents a systematic evaluation of existing and novel approaches for the statistical analysis of a series of N-of-1 trials. We derive practical recommendations which methods may be best in which scenarios.

## 1 Introduction

Within the last decade, personalized medicine has been on the rise. Treating patients on an individual level has been improved by the numerous possibilities to measure health outcomes with smart devices and application of novel data science approaches. In order to evaluate the effectiveness of health interventions on an individual level, N-of-1 trials have been established as the gold standard Nikles and Mitchell [2015], Davidson et al. [2021]. N-of-1 trials are multi-crossover controlled trials, where each patient is their own control group. In addition to individual-level analyses for personalized treatment, series of N-of-1 trials can be analyzed jointly Stunnenberg et al. [2018], or also combined with results from standard randomized controlled trials (RCTs) to obtain population-level estimates on the effectiveness of treatments with equal or superior efficiency compared to non-crossover RCTs Punja et al. [2016], Blackston et al. [2019]. In addition to research on appropriate statistical models for the analysis of aggregate and individual n-of-1 trials, previous studies have investigated approaches to derive optimal designs regarding sample size and number of cycles Diaz [2021], Senn [2019], Yang et al. [2021]. For the aggregate statistical analysis of series of N-of-1 trials, popularly used methods include non-parametric methods like the Wilcoxon signed-rank test Green et al. [2004], Sierra-Arango et al. [2019], two-sample mean tests Schmid CH, Duan N., the DEcIDE Methods Center N-of-1 Guidance Panel [2014], methods that allow for covariate adjustments like linear models Odineal et al. [2019], Vrinten et al. [2015], linear mixed models Herrett et al. [2021], and Bayesian approaches Chen and Chen [2014], Samuel et al. [2019]. Also, autoregressive models to account for time dependencies have been proposed for the analysis Zhou et al. [2017]. Daza introduced a counterfactual framework for time-dependent treatments to estimate average period treatment effects in N-of-1 trials Daza [2018]. This framework is also applicable to the analysis of n-of-1 observational studies, where the order of the treatment phases is not randomized and may be affected by confounding Daza and Schneider [2022].

Some studies have evaluated and compared different methods for the analysis. For example, Stunnenberg et al. Stunnenberg et al. [2018] applied both frequentist linear mixed models as well as Bayesian models, and compared the approaches in a study on the effect of mexiletine on muscle stiffness in patients with nondystrophic myotonia. Zucker et al. Zucker et al. [2010] compared repeated-measure models, Bayesian hierarchical models, and simpler single-period, single-pair, and averaged outcome crossover models in the analysis of a published series of N-of-1 trials on rheumatolic treatments. Their results showed that depending on the assumptions, different mixed models yielded the best fit and that Bayesian models were sensitive to the specification of the priors. Chen & Chen Chen and Chen [2014] compared t-tests and mixed models in a simulation study when no carry-over was present, and found t-tests to yield highest power under this assumption. Finally, Araujo et al. Araujo et al. [2016] extended the work of Chen & Chen and considered t-tests and linear mixed models under different model assumptions on the study design, with a focus on how the study design incorporated randomization.

In this study, we focus on two particular challenges for the analysis of aggregated N-of-1 trials: (i) carry-over and (ii) complex dependencies and time-varying interactions of the treatment effect with covariates. First, as a patient cannot receive two treatments at the same time, the treatment is time-varying. This may introduce carry-over- i.e., that the effect of one treatment is still active when the other treatment is applied - complicating the analysis of the trials and the interpretation of the results. As one solution, wash-out periods can be introduced in the study design, where the patient does not receive any of the treatments. However, this is not always possible, so statistical methods have to be investigated regarding their robustness against known or unknown carry-over. Second, for the aggregated analysis of N-of-1 trials, treatment effects may often depend on covariates, their effect might be modified by them, and this might be further complicated if time-varying interactions between treatment and effect modifiers exist. Carry-over and such complex dependencies have to be considered to ensure unbiased estimates of the causal treatment effects, but best-practise recommendations are not available Gamble et al. [2017].

Our paper is organized as follows. In Section 2, we describe a general data generation model and how we applied it to generate data for our simulation study. Then, we describe the evaluated statistical models which include our newly proposed carry-over adjusted parametric model (COAPM). In Section 3, we describe the results of the simulation study evaluating the performance of these statistical methods across four scenarios. We conclude with a discussion in Section 4.

## 2 Methods

In the following, we investigate different statistical methods for the analysis of aggregated N-of-1 trials on simulated data sets that contain different levels of carry-over and treatment-covariate dependencies. Additionally, we compare the methods on data sets with and without missing data due to participants’ dropout. As traditional methods, we include a sample mean comparison and linear regression model in the analysis Schmid CH, Duan N., the DEcIDE Methods Center N-of-1 Guidance Panel [2014], Nikles and Mitchell [2015]. Further, we introduce a parametric model that specifically models how carry-over modifies the treatment effect. Finally, we consider Bayesian Networks Pearl et al. [2016] and G-estimation Hernan and Robins [2019], Daza [2018].

To evaluate and compare the different statistical methods, we perform a Monte Carlo simulation study. In the following, we describe the simulation study set-up including the data generation model, a specific application of the data generation model to generate synthetic data of a series of N-of-1 trials on Chronic Nonspecific Low Back Pain, and the different evaluated statistical methods. The data generation model is available through the Python package sinot (github.com/HIAlab/sinot), and the statistical methods are implemented in the R package cinof1 (github.com/HIAlab/cinof1).

### 2.1 Data Generation

#### 2.1.1 Data Model

In the simulation model, we combine data generation based on stochastic processes, time-varying and covariate-dependent treatment effects, and effects onto the outcome variable embedded in a causal graph. For the notation in the following, let *Z* denote any of the variables in our model including the outcome *O*, treatment *T*, or other variables *C*.

First, we embed the outcome variable *O* and treatment variable *T* in a directed acyclic graph (DAG) with further variables *C*, which may be constant or time-varying, and can be simulated from several common distributions like Bernoulli, Gaussian, Poisson, or uniform. Time-invariant variables would not change over time and describe for instance Demographics or baseline conditions like Previous Diagnosis (of Nonspecific Low Back Pain). Time-varying variables may change on each observation and could be measurements like the number of steps per day.

We include linear effects from variables *Z*_*j*_ on *Z*_*i*_ at time point *t, t* ≥ 0, where *i* and *j* index distinct variables:

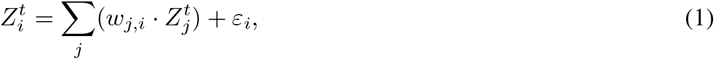

where *w*_*j,i*_ denotes the linear causal effect of *Z*_*j*_ on *Z*_*i*_ and *ε*_*i*_ denotes some random noise with mean *μ*_*i*_ and variance 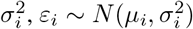. A treatment period may consist of one or more time points, i.e. days in our simulation.

To simulate binary variables, we define a threshold *λ*_*i*_ ∈ R and apply a step function *f* defined as:

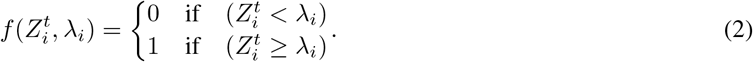

Time dependencies can be added to the data simulation by letting the variable 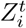 depend on the weighted values of *Z*_*j*_ at time points (*t* − *l*), i.e. adding lags *l*:

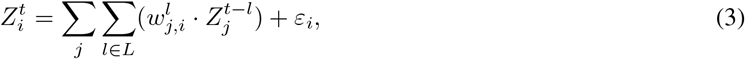

where *L* is a nonempty set of integers greater or equal to 0 and smaller or equal to *t*.

The treatment variables *T* have an exponential decay defined through wash-in *τ* and wash-out *γ* to simulate carry-over, similar to Percha et al. Percha et al. [2019] (see Supplementary Figure FS1 for an illustration). Daza described the carry-over as slow onset and slow decay Daza [2018].

After drawing the exogenous variables from pre-specified distributions, the endogenous variables are generated based on assumed weights *w*_*j,i*_ and the DAG.

To simulate the outcome *O*, we model an underlying state *U* with a baseline drift as a discrete-time stochastic process (Wiener process); see Supplementary Text 1 and Supplementary Figure FS2 for more details. Baseline drift here describes the observed change over time in the outcome variable if left untreated, which can be a time trend in specific cases (see Supplemental Text 1 for details). Then, *O* at time *t* is a linear combination of the causal effects of the other variables and the underlying state:

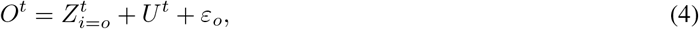

where 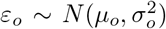, *U*^*t*^ denotes the underlying state at time point *t* and 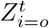 denotes the linear causal effects defined in the DAG of all covariates on the outcome variable *O* at time point *t* as defined in equation 3.

#### 2.1.2 Series of simulated N-of-1 trials of Chronic Nonspecific Low Back Pain

For the simulation study, our aim is to generate a realistic synthetic data set from a series of N-of-1 trials, comparing the effect of daily exercises for back strength (*Treatment 1*) with the effect of daily exercises for back stretching (*Treatment 2*) on reducing the outcome variable *Chronic Nonspecific Low Back Pain*. We assume that Pain is measured daily. The study includes two blocks of two treatment periods each.

We set each treatment period to a length of 4 weeks. The period order is randomly selected within each treatment block. An exemplary study scheme could look like *ABBA* or *ABAB*, where *AB* (or *BA*) would be a block with two treatment periods and a total study duration of 4x4 = 16 weeks. Additionally, the study contains a baseline assessment of medication use and different sociodemographic variables. We identified these variables on a literature review including Burdorf and Sorock [1997] and expert interviews.

*Demographics* encompasses variables gender and age at baseline, which are modeled as constant variables across all time points. *Education* assesses if the patient had an academic degree or was enrolled in an academic program. The variable *Work* identifies whether the patient is working or not. Both Work and Education are assumed as constant across all time points. Health status is assessed including daily measurements of *Medication*, (which identifies if a patient was using painkillers), *Previous Diagnosis* (of Nonspecific Low Back Pain), and *Chronic Diseases* (indicating whether a patient has been diagnosed with related chronic diseases, for instance scoliosis or muscular disorders). Besides health status, lifestyle factors are tracked on a daily basis, including physical *Activity, Stress* levels, and *Quality of Sleep*.

We create a DAG with the assumed causal relationships between all the identified variables, see Figure 1, based on a literature review and expert knowledge. We assume effects from Demographics on Education, Activity, Work, Previous Diagnosis, Medication and Chronic Diseases. Furthermore, we assume that there is no direct causal effect of Demographics on Nonspecific Low Back Pain, but an effect mediated by proxy variables, which leads to indirect causal paths from Demographics to Nonspecific Low Back Pain through, e.g., Activity. We assume that Treatment has an effect on Stress, Quality of Sleep, and Nonspecific Low Back Pain. We assume that Treatment does not affect Demographics, Education, Work, Chronic Diseases, Medication, and Previous Diagnosis, as they are assumed to be constant over time. As will be described in more detail in the next Section 2.1.3, we model a complex dependence of the treatment effect on Activity (see Figure 3).

**Figure 1:**
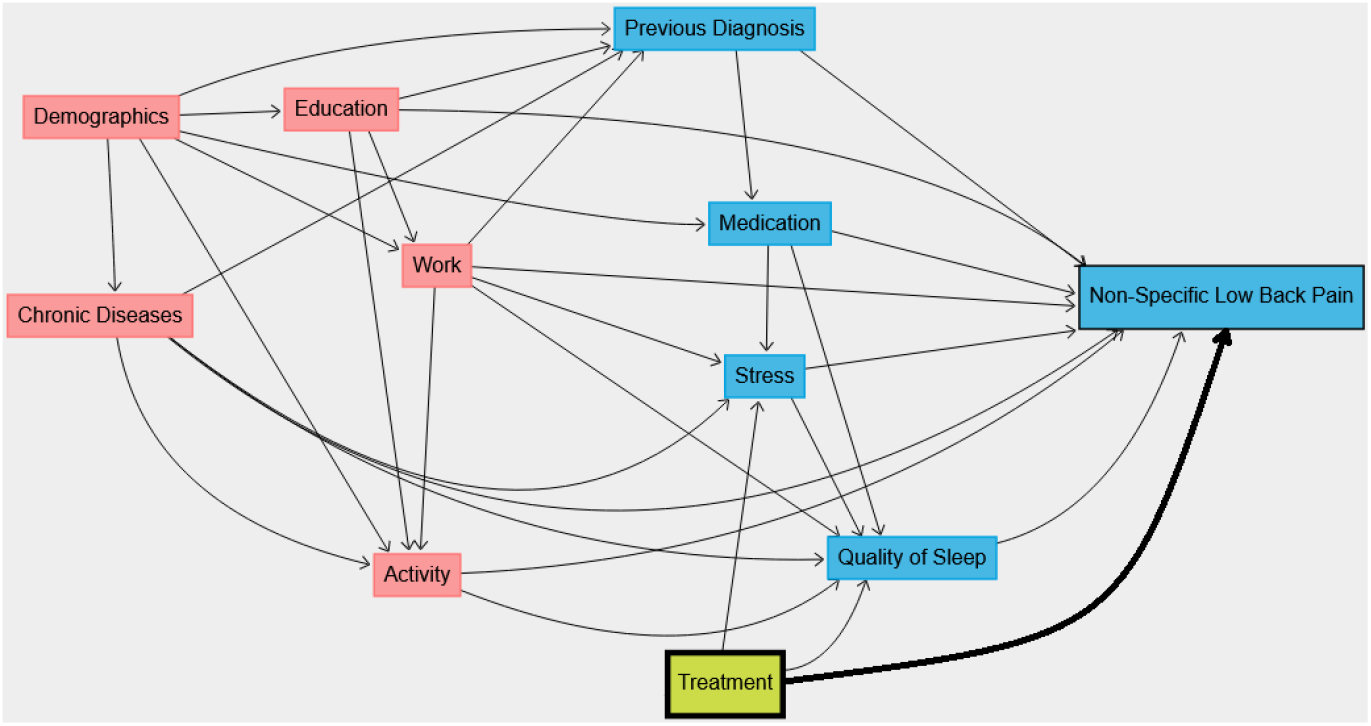
DAG of assumed causal effects in the simulated study. Variables having a direct or indirect effect on Activity were highlighted in red boxes as Activity is an interaction term in scenario 3. The Treatment variable is highlighted in green.

These effects are summarized in the DAG shown in Figure 1, and are used to generate the data. In addition to the dependencies shown in the graph, time dependencies are specified: Quality of Sleep as well as Activity depend on Treatment at the previous timepoint.

#### 2.1.3 Generated data sets

Based on the data generation model, the Nonspecific Low Back Pain study design, the DAG shown in Figure 1, and the time dependencies between variables described above, we generate data sets under four different scenarios, shown in Table 1.

**Table 1:** Overview of the different scenarios. In the scenarios with carry-over, the parameters for wash-in *τ* and wash-out *γ* simulating the exponential decay are changed. *w*_*A,T*_ denotes an effect modifier of Activity on Treatment. Hence, the treatment effect depends on Activity whenever *w*_*A,T*_ *≠* 0.

|  | No Activity interaction | Activity interaction |
| --- | --- | --- |
| **No carry-over** | *Scenario 1* | *Scenario 3* |
| | $\gamma = \tau = 1$ | $\gamma = \tau = 1$ |
| | $w_{A,T} = 0$ | $w_{A,T} \neq 0$ |
| **With carry-over** | *Scenario 2* | *Scenario 4* |
| | $\gamma_1 = 3; \tau_1 = 6$ | $\gamma_1 = 3; \tau_1 = 6$ |
| | $\gamma_2 = 4; \tau_2 = 5$ | $\gamma_2 = 4; \tau_2 = 5$ |
| | $w_{A,T} = 0$ | $w_{A,T} \neq 0$ |

All scenarios include covariate effects following the DAG in Figure 1, with the exception of the interactions and temporal effects between Activity and Treatment, which are only included in some scenarios (i.e., 3 and 4). In scenario 1, we generate the data as the baseline data set for all methods and do not include carry-over, time dependencies, or interaction between Activity and Treatment. In scenario 2, the data are simulated with carry-over so that the treatment effect is heavily time-dependent within a treatment period as we add wash-in and wash-out phases.

In scenario 3, a complex dependence of the Treatment effect on Physical Activity is modeled in the simulation additionally to the covariables shown in Figure 1. The observed Treatment variable indicating whether the patient is exposed to the Treatment or not is not affected by Physical Activity, but the underlying treatment effect is modified through *w*_*A,T*_. With that, we have modeled an interaction of Treatment and Activity. In addition, there is a temporal effect of Treatment at *t* − *l* on Activity at *t* so that the Activity distribution differs between the treatment groups. In scenario 4, we generate a data set with both carry-over and Treatment-Activity interactions as in scenario 3.

If the edge from a variable *j* to a variable *i* is present in the DAG, the effect is set to *w*_*j,i*_ *≠* 0. If the edge is not present in the DAG, it represents our assumption that there is no effect of *Z*_*j*_ on *Z*_*i*_; equivalently *w*_*j,i*_ = 0. Time dependencies are simulated in the same way, where we set 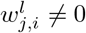 when we assume a time dependency between the variable 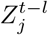 and the variable 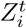. For all scenarios, the effects are identical except for the specifications mentioned in Table 1. Treatment effects were set to be constant over time. The effect of treatment 1 on the outcome was set to -2, and of treatment 2 on the outcome to -4, both compared to no treatment (i.e. baseline drift and covariate effects). Hence, this results in a treatment effect difference of 2 between the treatments (see Supplementary Text 2 for more details).

In addition to these four scenarios, we further investigate how the methods perform on data sets with missing values by replicating the 4 scenarios with missing values. For this, we use the same parameters and introduce row-wise missing values (i.e., across all variables of an individual) through two mechanisms. The first mechanism deletes 10% of the data points randomly with increased probability over time to mimic random drop-out. Second, we add a block of 10 consecutive missing days that were drawn randomly for each patient to simulate vacation (see Supplementary Text 3 for more details).

### 2.2 Statistical Methods

#### 2.2.1 Overview

As described in the previous section, we generate data sets from 4 different scenarios, each a series of N-of-1 trials on Chronic Nonspecific Low Back Pain with 1000 participants. For the evaluation of the statistical models, in each scenario, we draw 100 samples each of 5, 10, 25, 50 and 100 participants, to also investigate the influence of the sample size in aggregated N-of-1 trials. Then we apply different statistical models, and evaluate their bias and efficiency in estimating the treatment effect difference in outcomes between treatment groups 1 and 2 across all participants. We compare standard statistical models for the analysis of aggregated N-of-1 trials, COAPM, G-estimation, and Bayesian Networks.

#### 2.2.2 Standard Statistical Models

First, we compute the sample means of both treatment groups, and the naive estimate of the treatment effect difference. We call this the *Sample Mean* model. Its standard error is estimated as the empirical standard deviation of the estimated treatment effect difference across the 100 samples.

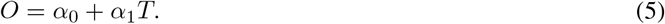

Second, we fit a standard multiple linear regression model with pain as the response variable, and the treatment and covariates as predictors. We call this the *Linear Model*. Hence, 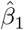 in model (6) is an estimate of the average direct effect of the treatment *T* on the pain outcome *O* adjusted for all covariates *C* in Figure 1 with direct effects on the outcome (i.e., excluding Demographics). That is, 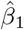 is an estimate of the direct effect of *T* on *O*:

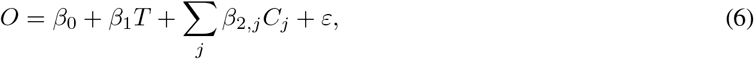

where *ε* follows a normal distribution. For the implementation, we use the `lm` function in `R` from the `base` package with default settings, assuming independence between observations of different patients, to compute non-weighted ordinary least squares estimates of the regression coefficients, along with standard error estimates and Wald test results. The effect estimates are then averaged across the 100 samples as the empirical mean, and standard error estimates are estimated as the empirical standard deviation of the effect estimates.

#### 2.2.3 Linear Models Adjusting for Wash-In and Wash-Out

To reduce the bias due to carry-over, we adjust the multiple linear regression model for wash-in *τ*_*k*_ and wash-out *γ*_*k*_, where *k* = 1 indicates treatment 1 and *k* = 2 indicates treatment 2. For that, we include a continuous time-dependent treatment effect variable instead of the binary treatment assignment variable. We call this the *carry-over adjusted parametric model (COAPM)*.

Let 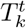 indicate whether the patient was exposed to treatment *k* at time point *t* and let 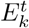 denote the exponential decay treatment indicator, which we parameterize given *τ*_*k*_ and *γ*_*k*_:

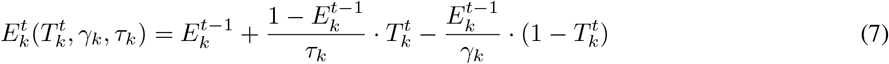

for *t* ≥ 1. We initialize 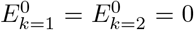, as we assume no treatment effect at the start point of the study. For an example, consider treatment *k* = 1 and wash-in *τ*_1_ = 2. Then, 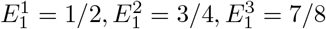…, That is, instead of using a treatment indicator *T* which takes values 0 or 1, *E*_*k*_ is a treatment indicator which incorporates wash-in and wash-out through exponential decay and either targets the value 1 (for wash-in) or 0 (for wash-out). Then we estimate the average effect of each treatment over time using the following linear regression model:

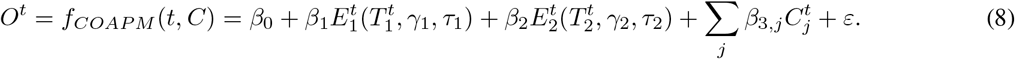

With that, 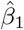 estimates the carry-over adjusted average effect of treatment 1 compared to no treatment (i.e. neither treatment 1 nor treatment 2, which would be baseline), and 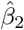 the carry-over adjusted average effect of treatment 2 compared to no treatment. Compared to the model in Equation 6, the COAPM model estimates the effect of 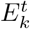, yielding an estimate of the carry-over adjusted treatment effect instead of the treatment indicator variable *T*_*k*_. Hence 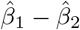 is an estimate of the treatment effect difference adjusted for carry-over.

As *E*_*k*_ is a function on *τ*_*k*_ and *γ*_*k*_ which are unknown, it is approximated through a grid search. In more detail, we iterate over several combinations of *τ*_*k*_ and *γ*_*k*_ and fit a linear model for each combination. Then, we estimate *τ*_*k*_ and *γ*_*k*_ from the model with highest *R*^2^ value. Estimates of *β*_1_ and its standard error are obtained from the final model with highest *R*^2^, using the `lm` function in `R` with default settings. The effect estimates and standard error estimates are then averaged across the 100 samples.

For an illustration, Figure 2 shows the treatment effects for two treatments with carry-over and the resulting overall treatment effect, which can be computed as the sum of the two treatment effects 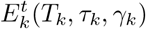. As the observed overall treatment effect contains the effects from both treatments, it over- or underestimates the treatment effects.

**Figure 2:**
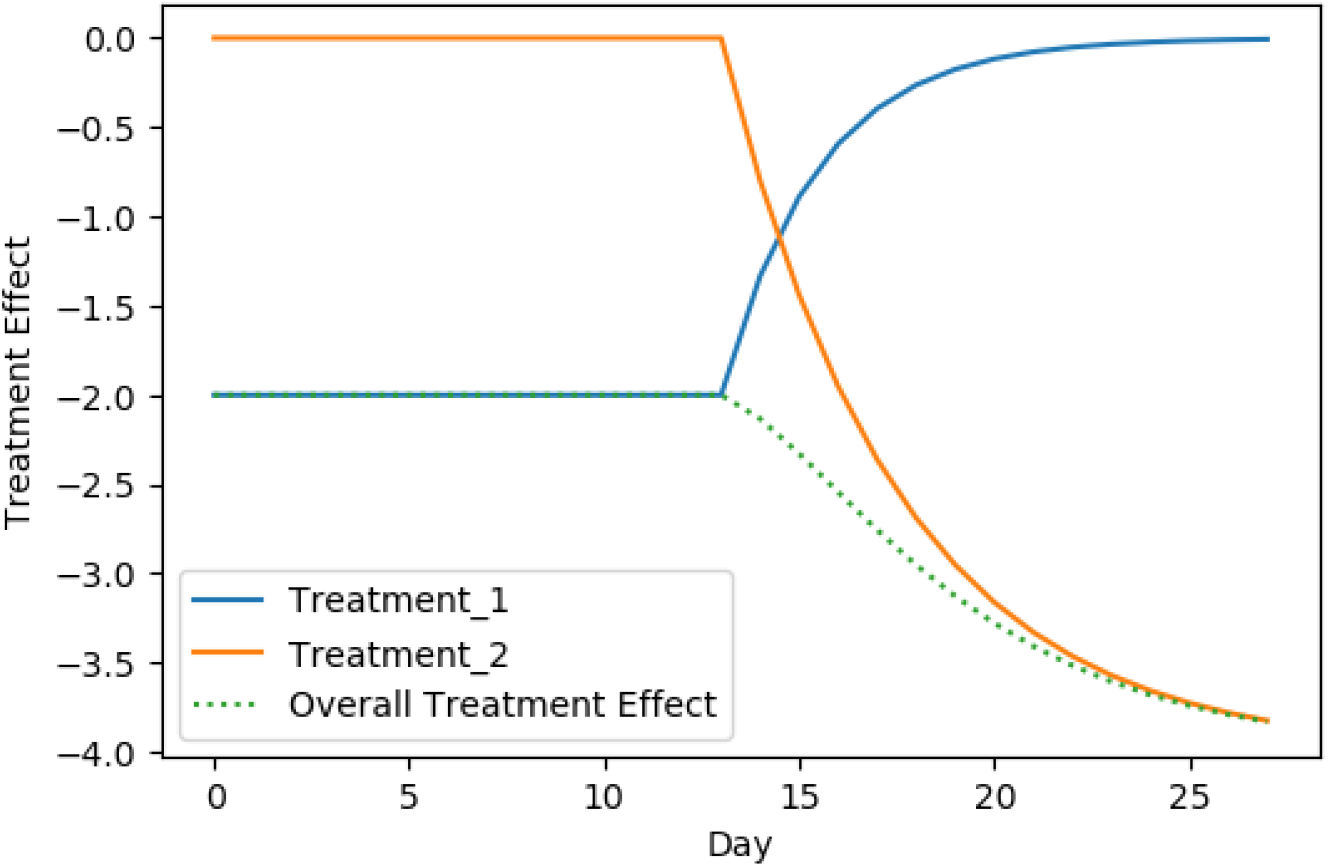
Illustration of the treatment effects with carry-over for a simulated patient. The patient was exposed to treatment 1 until day 14. Starting on day 14, the treatment effect of treatment 1 washes out and converges to 0 as the treatment was not given anymore. At this point, the patient started the second treatment period. With being exposed to of treatment 2, the effect of treatment 2 washes in and it takes time until it reaches the full effect on the outcome.

**Figure 3:**
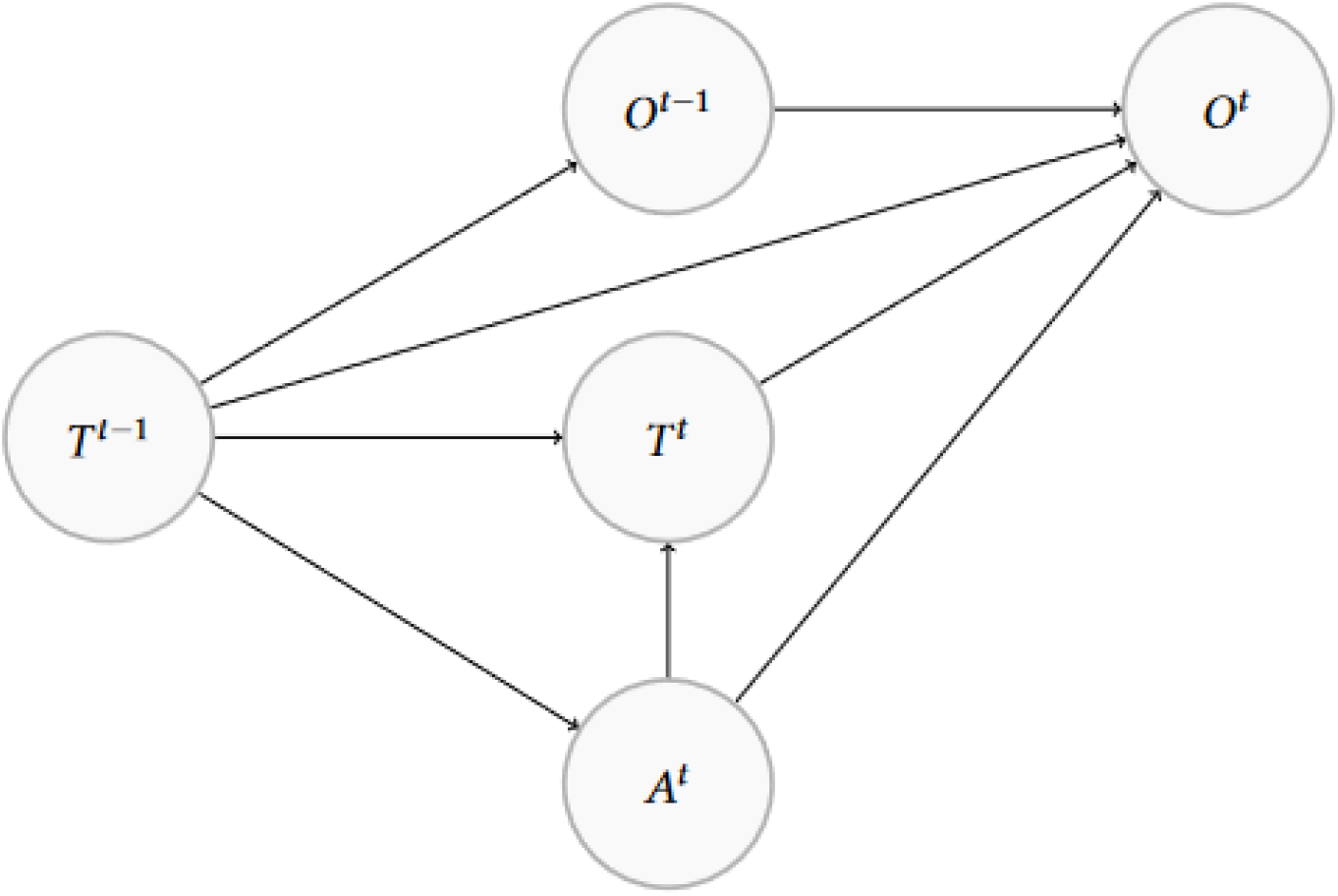
The DAG shows the assumed network structure between the nodes Activity (*A*), Treatment (*T*) and Nonspecific Low Back Pain (*O*) at time point *t* that is modeled in the Bayesian Network with time adjustment with lags. With that, the model is accounting for Activity as a confounding variable, although we do not simulate it. This is a snippet of the relationships between these three variables, and embedded in the bigger network in Figure 1.

#### 2.2.4 Bayesian Networks with Time-Dependent Variables

Bayesian Networks are graphical networks in which the joint and conditional probability distributions given by the assumed DAG are estimated, and can be used for inference. We call this the *unadjusted Bayesian Network model*. To consider time dependencies, we specify lags of size 1 for the treatment variable and for Nonspecific Low Back Pain (see Figure 3). These lags are created during preprocessing and are included in the analysis as additional variables. We call this the *Bayesian Network with time adjustment* model.

For the implementation, the `bnlearn` package is used. In the first step, we implement an interface to convert the `DAGitty` graph to a Bayesian Network and defined the appropriate scale of each variable. Then, the Bayesian Network is fitted to the data with the `bnlearn::bn.fit` function with default settings. We estimate the parameters by the empirical mean of their posterior distribution using the `method = “bayes”` argument. For estimating the average treatment effect difference, we use the `bnlearn::cpdist` function of the fitted network Scutari et al. [2017], and first generate two random samples of patients under treatment 1 and under treatment 2, each of size 1000 through likelihood weighting given the treatment and confounding variables to have equal confounding distributions among the treatment groups. Then, we estimate the average treatment effect difference by the mean outcome difference between the two random samples. The standard error estimate of the estimated treatment effect difference is calculated as the standard deviation of the estimated average treatment effect across the 100 samples.

In the analysis, we apply two Bayesian Networks. The first model is fitted to the DAG without any time dependencies. The second model additionally includes the lags to model time-dependencies as described above.

#### 2.2.5 G-estimation

G-estimation estimates average treatment effects - in our case, treatment effect differences in average outcomes between treatment groups - in a structural nested mean model Naimi et al. [2016], Hernan and Robins [2019], and can be applied to both time-varying and time-invariant treatment variables. Rubin [1974], Holland [1986], Splawa-Neyman et al. [1990]

In the structural nested mean model, following the notation of Hernán and Robins (2019), we model the expected conditional difference between the outcome under treatment 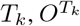, and the potential outcome under the first treatment, 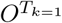, as

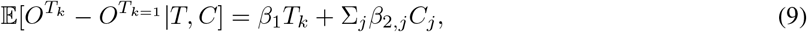

where *β*_1_ denotes the average treatment, *C* includes all variables (with direct or indirect effect on the outcome) observed in the data set, and *T*_*k*_ indicates whether treatment 1 (*T*_*k*=1_) or treatment 2 (*T*_*k*=2_) was given. For estimating the average treatment effect difference using G-estimation, we search for *ψ* which minimizes |*θ*_1_| in the following equation:

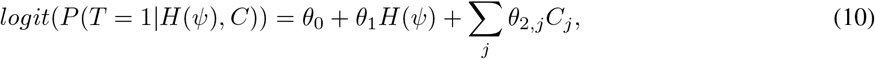

where *ψ* is the individual causal effect induced by the corresponding assumed rank-preserving model and *H*(*ψ*) is defined as *H*(*ψ*) = *O* − *ψT*. We assume that the conditional exchangeability assumption holds, which implies that |*θ*_1_| should be 0 at the true *ψ*. In this way, minimizing |*θ*_1_| allows us to estimate the true *ψ*. We assume that conditional additive rank preservation holds, such that *ψ* = *β*_1_, the average treatment effect of interest.

Compared to the Bayesian Network with time adjustment and the unadjusted Bayesian Network, we do not use lags in this model. Here, we used generalized estimating equations with *independence* and autoregressive order 1 (*AR1*) working correlation structure from the R-package `geepack` Højsgaard et al. [2005] to fit equation 10. We call these the *G-estimation (independence)* and *G-estimation (AR1)* models, respectively. In the model based on the AR1 correlation matrix in the generalized estimating equations, the value for the estimated effect difference is based on the difference between the treatment effects incorporating the introduced wash-in. The standard error estimate of the estimated treatment effect difference is calculated as the standard deviation of the estimated average treatment effect across the 100 samples.

## 3 Results

As described in section 2.1.3, we consider four different scenarios in the simulation study. In each scenario, all methods were evaluated on 100 samples of 5, 10, 25, 50, and 100 patients, respectively. Figure 4 shows the mean estimates of the treatment effect difference for all models, in all scenarios, with and without missing values, along with their respective standard error estimates. Supplementary Text 4 provides the plotted numeric values and other details.

**Figure 4:**
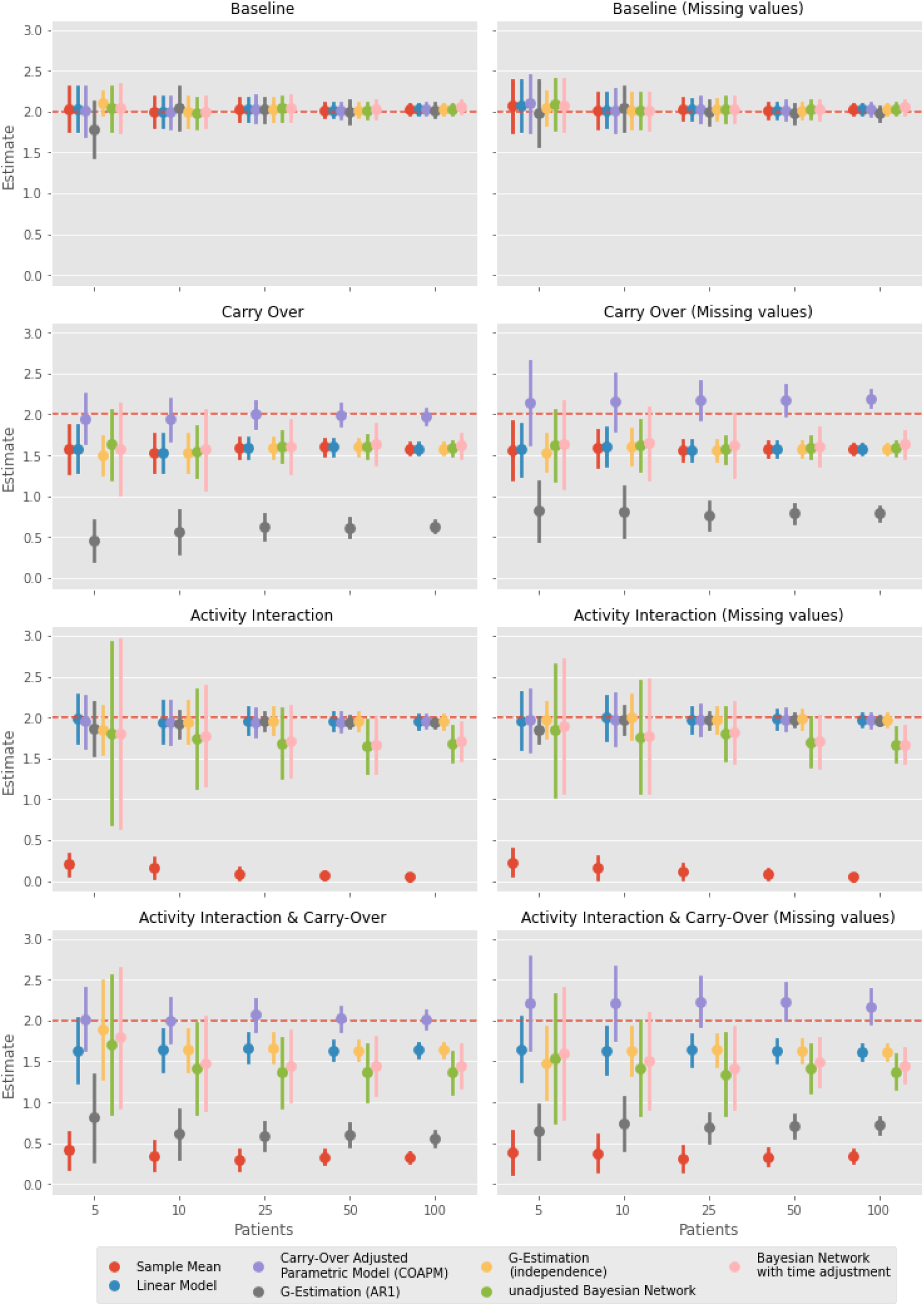
Overview of the estimates of the treatment effect differences (y-axis), with a true value of 2 (broken red horizontal line), with standard error bars, across the four scenarios (1-4 displayed in order from top to bottom) with and without missing values, for different sample sizes on the x-axis.

### 3.1 Scenario 1: No Carry-Over and No Activity interaction

In the first scenario without Activity interaction and without carry-over, all methods provide unbiased estimates of the true treatment effect difference of 2. As can be expected, the estimates are more efficient; i.e. had smaller standard errors for larger sample sizes. For the smallest sample size with 5 patients, the G-Estimation (AR1) model slightly underestimates the treatment effect as it assumes autocorrelation, which is not present in this scenario. For the data with missing values, the models also provided unbiased treatment effect estimates, with expected slightly larger standard errors as we have fewer observations.

### 3.2 Scenario 2: Carry-Over Only

In the second scenario, we investigate wash-in and a wash-out influences to our treatment variables. With these, the true simulated treatment effect slightly increases over time up to the full treatment effect for each treatment compared to no effect (i.e., zero). As a result, the sample mean model, linear model, G-estimation (independence), and the Bayesian Network with time adjustment all underestimate the simulated treatment effect difference of 2. As the treatment effect increases over time up to the full effect size, the models could not estimate the simulated treatment effect size. Compared to the unadjusted Bayesian Network, we expected the Bayesian Network with time adjustment to improve the estimated treatment effect difference, which was not reflected in the results as the model did not improve the estimate and also underestimated the treatment effect difference. G-estimation (AR1) assumes an AR1 dependence structure within the data, violated through the exponential decay. Hence, it strongly underestimates the treatment effect in simulation with carry over. The only model that yields unbiased estimates of the treatment effect difference for all sample sizes was the COAPM with parameters for wash-in and wash-out, which is close to the data simulation process.

Within the data set with missing values, the COAPM tends to overestimate the treatment effect. The other models performed similarly poorly (with respect to bias) when there were missing values compared to no missing values, but with slightly increased standard errors.

### 3.3 Scenario 3: Activity interaction Only

In this scenario, we are considering complex Treatment effect dependencies on Activity. As expected, the sample mean model heavily underestimates the treatment effect for all sample sizes. The linear model, COAPM, G-Estimation (AR1), and G-Estimation (independence) all provide unbiased estimates for all sample sizes. Surprisingly, both the Bayesian Network with time adjustment and the unadjusted Bayesian Network slightly underestimate the treatment effect difference, which becomes more apparent for larger sample sizes. It could be, that the priors were uninformative in this scenario or the number of cycles was too small. As another observation, both the Bayesian Network with time adjustment and the unadjusted Bayesian Network yield larger standard errors compared to linear models. This could be due to the fact, that we model Activity as an effect modifier and with temporal dependencies. By increasing the sample size, all standard errors decrease. In this scenario, missing values yield slightly larger standard errors.

### 3.4 Scenario 4: Carry-over and Activity interaction

The last investigated scenario contains both carry-over and Activity interaction. We observe that both the sample mean and G-estimation (AR1) strongly underestimate the treatment effect, both for complete data and data with missing values. Both the Bayesian Network with time adjustment and unadjusted Bayesian Network provide treatment effect difference estimates of about 1.5, hence underestimating the effect difference, and also yield larger standard errors compared to the other methods as seen already in scenario 3. The Bayesian Network with time adjustment yields slightly better results than the unadjusted Bayesian Network, but does not provide a major improvement. The linear model and G-estimation (independence) provide less biased treatment effect estimates, but still also underestimate the treatment effect difference. Finally, the COAPM again provides good results in this scenario. Across all sample sizes with complete data, this model yields unbiased estimates of the treatment effect difference. When data points are missing, this model slightly overestimates the treatment effect.

### 3.5 Summary

Overall, COAPM yields robust results, the best results among all considered models, in all scenarios with complete data. However, when data is missing and carry-over is present, then this approach tends to overestimate the treatment effect difference. Linear models and 2-sample t-tests are robust against simulated missing values, but yield biased effect estimates when strong carry-over is present as they are not adjusted for it. Furthermore, the sample mean yields biased effect estimates when Activity interaction is present. Bayesian Networks and G-estimation show a good overall performance, but Bayesian Networks yield wider confidence intervals of effect estimates especially for small sample sizes.

## 4 Discussion

In this study, as a first contribution, we demonstrate how to simulate data for a series of N-of-1 trials by marrying stochastic processes with time-varying treatment effects embedded in a DAG. In our complex simulation models, we made assumptions about the causal structure underlying Chronic Nonspecific Low Back Pain, and provide recommendations for analyses, that can be translated into an actual conducted series of N-of-1 trials. As a main contribution, we evaluate and compare different models for estimating the treatment effect under the presence of carry-over, complex dependencies of the treatment effect on covariates, and missing values. These results can provide guidelines which methods should be used in practical applications, and we provide the R package cinof1 (available from github.com/HIAlab/cinof1) with an implementation of all investigated methods.

One of our main findings is that simple statistical models can provide unbiased treatment estimates across different scenarios. Furthermore, we show that if carry-over is present and has not been prevented by the study design (e.g. by including wash-out phases), it is possible to still obtain unbiased treatment effect estimates when the carry-over is modeled in the analysis. This adds an interesting novel perspective, in contrast to previous studies which have largely focused on the removal of carry-over through study design, and have recommended against the adjustment for carry-over in statistical modeling Araujo et al. [2016]. For this situation, we provide a simple method called COAPM to incorporate carry-over into linear regression models. COAPM yields unbiased estimates even if a strong carry-over is present, but requires complete data. Finally, our results showed that G-estimation and both the Bayesian Network with time adjustment and the unadjusted Bayesian Network can provide unbiased and efficient treatment estimates, but they suffer from limitations in some scenarios.

Simple methods like sample mean comparisons and linear models are easy to apply and evaluate. They are also robust to missing values and applicable for any sample size, and deliver good results on the data sets without strong carry-over and without treatment-activity dependencies. This is in line with the results from previous studies that t-tests yield robust and valid results Chen and Chen [2014], Araujo et al. [2016]. On the other hand, sample mean comparisons do not account for carry-over and time dependencies, and did not yield good results in the presence of confounding. Linear models performed better, but do not account for carry-over.

In order to model carry-over, we introduce COAPM for wash-in and wash-out. It yields unbiased estimates for the treatment effects difference across all data sets, except for some scenarios with missing data. Here, it overestimates the treatment effects but still has less bias than all other methods. All other investigated methods are not able to yield unbiased treatment effect estimates when there is carry-over. As the COAPM was close to the data simulation, it delivered the best results across the different scenarios.

G-estimation performed very similar to linear models unadjusted for carry-over, yielding unbiased treatment effect estimates across many scenarios when there is no carry-over, and is robust to missing values. However, how the GEE correlation structure is specified proved to be very important, and AR1 yields largely biased estimates when there is carry-over as wash-in and wash-out periods are simulated through an exponential decay and not as an AR1 process. This was interesting to observe as it could be hypothesized that even a misspecified AR1 working correlation can make the GEE estimator more statistically efficient. But this was not observed in the results, so it seems that the misspecification played a larger role and the small sample size might have also contributed.

Finally, we investigated two implementations of both the Bayesian Network with time adjustment and the unadjusted Bayesian Network which show robust results with respect to variations in sample size and missing values when there is no carry-over, similar to G-Estimation. Interestingly, the Bayesian Network with time adjustment did not outperform the unadjusted Bayesian Network. In the Bayesian Network with time adjustment, we included lags of 1 in the network. However, the exponential decay used in the simulation takes multiple previous states of the treatment into account, which are not reflected in the model. We hypothesize that this misspecification of the time dependency led to this model’s poor performance. For fitting Bayesian Networks, a graph has to be constructed in a first step. This can be computed based on the data, but is not recommended [Hernan and Robins, 2019, Chapter 6.5]. Assuming a pre-specified DAG is preferred for interpretability, similar to all other investigated methods. Furthermore, the DAG serves to ensure generalizability, since it is not constructed on the sample data but a priori. It should be noted that we obtained parameter estimates from Bayesian Networks in order to compare the results to the other methods in this study; however, the full posterior distribution of the parameters are estimated in Bayesian Networks, allowing for other analyses and interpretations if desired.

One limitation of our simulation study is that we only included linear dependencies and fixed effects. In follow-up studies, nonlinear dependencies and random effects models could be incorporated to provide even more realistic data models. The Nonspecific Low Back Pain application that we considered provided a complex N-of-1 trial, and necessitated a complex generation of the DAG and simulation. For this study, we generated an outcome variable measured on an ordinal scale. In the analysis, however, we modeled the variable as a truncated Gaussian outcome. While this provides some model misspecification of all models that we investigated, we chose this evaluation to mimic a situation that occurs very often in practical analyses. In follow-up studies, other outcome distributions and other statistical models for the analysis can be investigated. Additionally, the study design, number of cycles, length of treatment periods, and baseline periods can affect the model performance, but were all not investigated in our study.

We also examined the impact of missing values. In practical applications, it is recommended to include some form of imputation, for example, multiple imputation. This would be especially important for the application of time-dependent methods and when the data are not missing completely at random.

In follow-up analyses, it would be interesting to compare these methods in a real series of N-of-1 trials on Chronic Nonspecific Low Back Pain. Furthermore, additional methods like propensity score matching and inverse probability weighting could be interesting for analyzing aggregated N-of-1 trials, especially when there is missing data and selection bias.

We plan to further develop the R package with all implemented methods to handle plausibility checks and include further automated tests, and to provide a computationally more efficient process of estimating *τ*_*j*_ and *γ*_*j*_ in the COAPM compared to the currently implemented grid search. Finally, we think that incorporating an adjustment for carry-over into G-estimation or Bayesian Networks, and investigating the use of autoregressive moving average models including exogenous covariates (ARIMAX), e.g. Daza and Schneider [2022]’s n-of-1 ARCO model, in addition to methods to control for selection bias, can provide even more powerful and robust tools to estimate causal treatment effects in series of N-of-1 trials.

## Data Availability

All data produced and analysed in the present study are available in the HIAlab/sinot repository on github.com

https://github.com/HIAlab/sinot

## Backmatter

### Author’s contributions

Conceptualization, T.G. and S.K.; methodology, T.G. and S.K.; formal analysis and data generation, T.G.; writing-original draft preparation, T.G.; writing-review and editing, S.K., J.S and T.G.; supervision, S.K. and B.A.

All authors read and agreed to the final version of the manuscript.

## Acknowledgements

The authors wish to thank Dr. Tilman Engel for critical feedback on chronic nonspecific low back pain for the generation of the directed acyclic graph.

## Availability of data and materials

The datasets generated and analyzed during the current study are available in the `HIAlab/sinot` repository at github.com/HIAlab/sinot. The R code and Python code used for the analysis in this study is available at GitHub:

- R package for the statistical analysis: github.com/HIAlab/cinof1
- Python package for the data simulation: github.com/HIAlab/sinot

## Supplemental material

The following supporting information are attached:

- Supplementary Text 1: Stochastic Time-Series Data Generation Model
- Supplementary Text 2: Details on simulation study on nonspecific low back pain
- Supplementary Text 3: Simulation of Missing Values
- Supplementary Text 4: COAPM Model
- Supplementary Text 5: Full Results of the Simulation Study

---

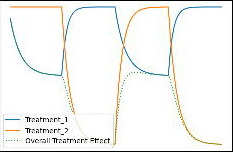

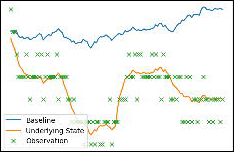

