## Supplemental Material for "Comparison of Bayesian Networks, G-estimation and linear models to estimate causal treatment effects in aggregated N-of-1 trials with carry-over effects"

Thomas Gärtner, Juliana Schneider, Bert Arnrich and Stefan Konigorski

### Supplementary Text 1. STOCHASTIC TIME-SERIES DATA GENERATION MODEL

In our data simulation model, we used a similar model as described in Percha et al. [1] for the stochastic time series component. Let $T$ denote the treatment variable, where $T_k(t) = 1$ indicates that treatment $k$ is given at time point $t$ and $T_k(t) = 0$ otherwise. We assume that the wash-in $\tau_k$ and wash-out $\gamma_k$ of the treatment effect can be described by the following ordinary differential equation:

$$dE_k = \left[\frac{1 - E_k}{\tau_k} T_k(t) - \frac{E_k}{\gamma_k}\left(1 - T_k(t)\right)\right] dt \tag{ES1}$$

This equation captures the decay of the wash-in and wash-out. Together with the linear effect of the treatment on the outcome, $w_{T_k,O}$, and by setting $Z^t_{T_k} = E^t_k$ in equation (3) in the main manuscript, the direct carry-over adjusted treatment effect of treatment $k$ at timepoint $t$ on the outcome is $w_{T_k,O} \cdot E^t_k$. In the simulation, we incorporated exponential decay in the modeling of the full treatment effect at discrete time points $t$. For that, we used equation ES1 as the difference between two time points:

$$\begin{aligned} E^t_k - E^{t-1}_k &= \frac{1 - E^{t-1}_k}{\tau_k} T_k(t) - \frac{E^{t-1}_k}{\gamma_k}\left(1 - T_k(t)\right) \\ E^t_k &= E^{t-1}_k + \frac{1 - E^{t-1}_k}{\tau_k} T_k(t) - \frac{E^{t-1}_k}{\gamma_k}\left(1 - T_k(t)\right) \end{aligned} \tag{ES2}$$

As a result, we have a recursive formula for generating the exponential decay. This also shows that $E^t_k$ can be understood as an exponential decay treatment indicator. Figure FS1 shows the simulated treatment effect with the exponential decay.

Further, a baseline drift $B$ depending on time $t$ is introduced through a Wiener process:

$$B(t + \Delta t) = B(t) + \Delta B(t) \tag{ES3}$$

where $\Delta B(t) \sim N(\mu_b, \theta_b)$ (see [1] for more details). With that, we can simulate positive or negative time trends, when we set $\mu \geq 0$ or $\mu \leq 0$, respectively.

Figures FS1 and FS2 show an illustration of the simulated data. In Figure FS1, the treatment effects $w_{T_k,O} E_k$ (blue and orange) and the overall treatment effect $\sum_k w_{T_k,O} E_k$ (green) are plotted. The patient has 4 treatment periods with a length of 28 days each and was exposed to $T_1, T_2, T_1, T_2$. Figure FS2 illustrates the different components of the outcome: the baseline drift (blue), the underlying state (orange) and the observed outcome values (green). In this example, the mean of the noise in the baseline drift $\mu_b$ was set to 0 so the baseline does not change over time. If $\mu_b$ were different from 0, then it would be expected that the baseline drift increases or decreases over time. For instance, if $\mu_b$ were set to -1, it would be expected that the outcome on day $i$ decreases compared to the previous day $\mathbb{E}(O_i) = \mathbb{E}(O_{i-1}) - 1$.

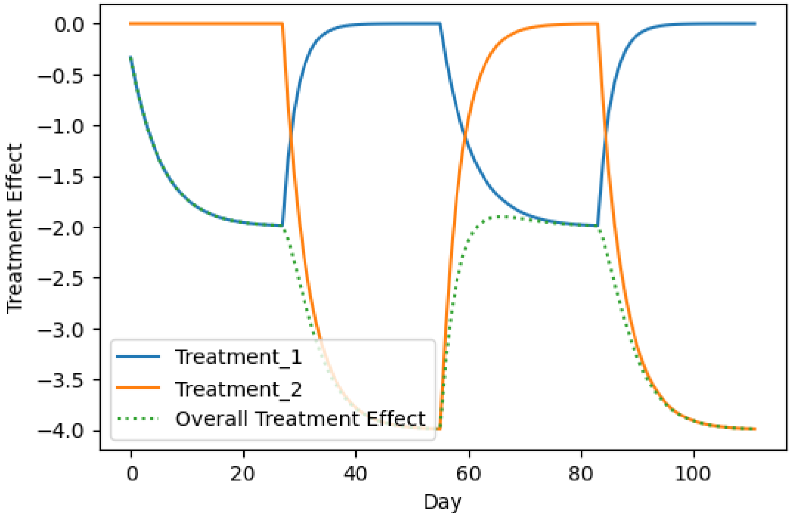


**Fig. FS1. Treatment Effect.** Treatment effects over time of 2 treatments with wash-in and wash-out effects and the overall treatment effect.

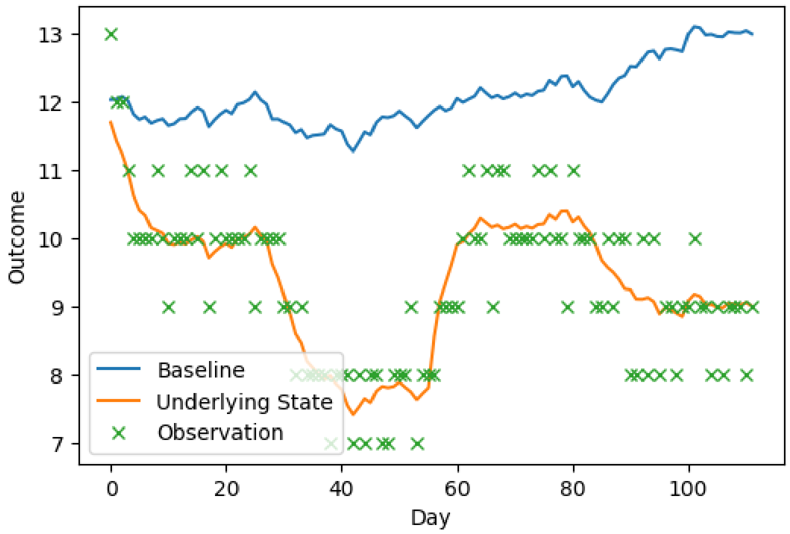


**Fig. FS2. Outcome Simulation.** Example outcome simulation with baseline drift ($\mu_b = 0$), underlying state and observations over time.

### Supplementary Text 2. DETAILS ON SIMULATION STUDY ON NONSPECIFIC LOW BACK PAIN

In the first step, we identified relevant variables as shown in Table TS1 and defined the variable measurements. The treatment was simulated as a binary variable identifying whether the patient was getting treatment 1 or 2, and had a certain effect size, wash-in effect and wash-out effect depending on the simulation scenario. All patients in the cohort were exposed to each treatment in each treatment block. We simulated a bigger effect for back stretching (treatment 2) in each data simulation by setting the effect from treatment 1 on the outcome to -2 and from treatment 2 on the outcome to -4.

The outcome variable nonspecific low back pain was assessed on discrete pain levels on a scale from 0 to 15, where 15 indicates highest pain and 0 indicates no pain, drawn from a normal distribution through the Wiener process. To ensure that the simulated variables were in the defined range and discrete, they were truncated and rounded, if applicable.

$$O_j = \begin{cases} 15 & \text{if} \quad 15 \leq O_j \\ O_j & \text{if} \quad 0 < O_j < 15 \\ 0 & \text{if} \quad 0 \geq O_j \end{cases} \tag{ES4}$$

As there are several scales to quantify the outcome [2], the simulated outcome can be interpreted as the score from the *low back pain rating scale* [2] divided by four. In all simulated data sets, the noise variable was generated with the same variance of 1. The mean $\mu_b$ of the baseline drift was set to 0 and the standard deviation $\theta_b$ to 0.2 to have an effect on the outcome over time. We assumed that all patients were between 20 and 60 years old (drawn uniformly) and had a balanced male-female distribution.

As a proxy for the activity of a patients, *Steps per Day (Activity)* were simulated from a normal distribution $N(\mu = 8000, \theta = 500)$ and the effects from Education, Demographics, Work and Chronic Diseases were added. The *Quality of Sleep* was measured on a scale of 0 (good quality) to 10 (bad quality), which could come from a questionnaire. The parameter was simulated with a truncated normal distribution $N(\mu = 5, \theta = 3)$. *Level of Stress* could come as well from a questionnaire where the patient quantify the stress on a scale from 0 (no stress) to 10 (high stress). For the simulation, a Poisson distribution with $\lambda = 2$ was used. See Table TS1 for more details on the parameter values.

**Table TS1.** Overview of the parameters used in the data simulation for the treatment, outcome and covariates. $U(lower, upper)$ denotes a uniform distribution with a lower and an upper bound, $\lambda$ is the decision parameter for the step function explained in the main manuscript and $N(\mu, \sigma)$ refers to a Normal distribution with corresponding values for the mean and the standard deviation. The simulation order of the variables in the data set matches the order of the variables in the table.

| Category | Variable Name | | Simulation Parameter Baseline |
|---|---|---|---|
| Treatment | Exercises for back strength | $E_1^t$ | $\gamma_{=}1; \tau = 1$ |
| | Exercises for back stretching | $E_2^t$ | $\gamma_{=}1; \tau = 1$ |
| Covariates | Demographics (Age) | $C_D$ | $= U(20, 60)$ |
| | Education | $C_E$ | $= U(0,1) - 0.02C_D \quad \vert\lambda = 0.1$ |
| | Chronic Diseases | $C_{CD}$ | $= U(0,1) + 0.01C_D \quad \vert\lambda = 0.5$ |
| | Previous Diagnosis | $C_{PD}$ | $= U(0,1) + 0.03C_D + 0.01C_E \quad \vert\lambda = 0.4$ |
| | Medication | $C_M$ | $= U(0,1) + 0.025C_D + 0.01C_{PD} \quad \vert\lambda = 0.2$ |
| | Work | $C_W$ | $= U(0,1) + 0.001C_D + 0.05C_E \quad \vert\lambda = 0.7$ |
| | Level of Stress | $C_{LS}$ | $= 0.5C_{CD} + 1C_M + 0.2C_W + Poisson(\lambda = 2)$ |
| | Steps per Day (Activity) | $C_A$ | $= 1000C_W + 0.01C_E - 100C_D - 1000C_{CD} + N(8000, 500)$ |
| | Quality of Sleep | $C_{QS}$ | $= 0.001C_A - 0.01C_{CD} - 0.5C_M + 0.4C_{LS} - 0.3C_W + N(5, 3)$ |
| Outcome | Chronic Nonspecific LBP | $O$ | $= B(t) - 2E_1^t - 4E_2^t - 0.0002C_A + 0.02C_{CD} + 0.3C_E - 0.1C_M$ $+ 0.5C_{PD} + 0.01C_{QS} + 1C_W + 0.2C_{LS} + N(0, 0.5)$ |

### Supplementary Text 3. SIMULATION OF MISSING VALUES

For simulating realistic data sets, missing values were introduced by randomly setting selected observations to *None*. The probability of setting a data point to missing was increased linearly over time so that the patient was more likely to have missing values as the trial progresses. With that, missing values are not completely at random, but as we have set the treatment order randomly we do not expect systematic bias on the treatment effect difference between the two treatments.

Furthermore, random blocks with missing values were added. These blocks had a predefined length and were inserted at a random time point with equal probability over time. In summary, for generating data with missing values, two parameters were specified in the simulation. The first parameter describes the dropout rate, which is the ratio of missing values to all data points. The second parameter defines the length of a block of missing values.

### Supplementary Text 4. COAPM MODEL

In our data generation model, we used the following equation:

$$O^t = Z^t_{i=o} + U^t + \varepsilon_o, \tag{ES5}$$

where $Z^t_i = \sum_j \sum_{l \in L}(w^l_{j,i} \cdot Z^{t-l}_j) + \varepsilon_i$. For simplicity, we set $L = [0]$ and substitute it in the $Z^t_{i=o}$:

$$O^t = \sum_j (w_{j,i} \cdot Z^t_j) + U^t + \varepsilon_o \tag{ES6}$$

In the next step, we extract the treatment variables from the sum, so that $j$ is a set of covariates $C$:

$$O^t = w_{T_1,O} \cdot Z^t_{T_1} + w_{T_2,O} \cdot Z^t_{T_2} + \sum_{j \in C}(w_{j,i} \cdot Z^t_j) + U^t + \varepsilon_o, \tag{ES7}$$

where $Z^t_{T_k} = E^t_k$:

$$O^t = w_{T_1,O} \cdot E^t_{T_1} + w_{T_2,O} \cdot E^t_{T_2} + \sum_{j \in C}(w_{j,i} \cdot Z^t_j) + U^t + \varepsilon_o. \tag{ES8}$$

In order to estimate $w_{T_1,O}$ and $w_{T_2,O}$, we used the COAPM model defined as:

$$f_{COAPM}(t, C) = \beta_0 + \beta_1 \cdot E^t_{T_1} + \beta_2 \cdot E^t_{T_2} + \sum_{j \in C}(\beta_j \cdot C) + \varepsilon. \tag{ES9}$$

Assuming that $\beta_0 = \mathbb{E}(U)$, we estimate $w_{T_k,O}$ with $\hat{\beta}_k$ in the COAPM model.

Also, we estimate the treatment effect difference with $\hat{\beta}_1 - \hat{\beta}_2$ .

### Supplementary Text 5. FULL RESULTS OF THE SIMULATION STUDY

For each scenario, 100 samples with 5, 10, 25, 50 and 100 patients were drawn and analyzed separately. Estimates of the mean ($\mu$) and standard error of the mean ($\sigma$) treatment effect difference were obtained using the different models.

**Table TS2. Scenario 1: Baseline.** Overview of the estimates (of the mean, $\hat{\mu}$, and standard error of the mean, $\hat{\sigma}$) for the different sample sizes for the models: carry-over adjusted parametric model (COAPM), unadjusted Bayesian Network (BN), Bayesian Network with time adjustment (BNt), G-Estimation - independence (GEST-i), G-Estimation - AR1 (GEST-AR1), Linear Model (LM), and Sample Mean (SM).

| **Missing** | **Patients** | 5 | | 10 | | 25 | | 50 | | 100 | |
|---|---|---|---|---|---|---|---|---|---|---|---|
| **Values** | **Method** | $\hat{\mu}$ | $\hat{\sigma}$ | $\hat{\mu}$ | $\hat{\sigma}$ | $\hat{\mu}$ | $\hat{\sigma}$ | $\hat{\mu}$ | $\hat{\sigma}$ | $\hat{\mu}$ | $\hat{\sigma}$ |
| **Without** | **COAPM** | 2.006 | 0.296 | 1.989 | 0.196 | 2.032 | 0.158 | 2.010 | 0.094 | 2.030 | 0.069 |
| | **BN** | 2.036 | 0.276 | 1.989 | 0.185 | 2.039 | 0.146 | 2.013 | 0.095 | 2.027 | 0.064 |
| | **BN-t** | 2.044 | 0.299 | 1.999 | 0.185 | 2.045 | 0.153 | 2.033 | 0.109 | 2.055 | 0.080 |
| | **GEST-i** | 2.106 | 0.143 | 1.999 | 0.180 | 2.032 | 0.137 | 2.018 | 0.087 | 2.031 | 0.059 |
| | **GEST-AR1** | 1.779 | 0.359 | 2.042 | 0.269 | 2.020 | 0.149 | 1.996 | 0.135 | 2.014 | 0.086 |
| | **LM** | 2.032 | 0.271 | 1.996 | 0.179 | 2.032 | 0.137 | 2.018 | 0.087 | 2.031 | 0.059 |
| | **SM** | 2.032 | 0.274 | 1.994 | 0.182 | 2.031 | 0.138 | 2.018 | 0.087 | 2.031 | 0.060 |
| **With** | **COAPM** | 2.102 | 0.348 | 2.014 | 0.258 | 2.026 | 0.156 | 2.018 | 0.115 | 2.028 | 0.081 |
| | **BN** | 2.085 | 0.312 | 2.013 | 0.219 | 2.026 | 0.152 | 2.025 | 0.107 | 2.034 | 0.073 |
| | **BN-t** | 2.080 | 0.319 | 2.006 | 0.229 | 2.022 | 0.152 | 2.024 | 0.114 | 2.050 | 0.088 |
| | **GEST-i** | 2.048 | 0.217 | 2.014 | 0.211 | 2.025 | 0.124 | 2.016 | 0.094 | 2.027 | 0.065 |
| | **GEST-AR1** | 1.977 | 0.428 | 2.036 | 0.266 | 1.992 | 0.149 | 1.980 | 0.115 | 1.974 | 0.081 |
| | **LM** | 2.076 | 0.305 | 2.013 | 0.209 | 2.025 | 0.124 | 2.016 | 0.094 | 2.027 | 0.065 |
| | **SM** | 2.067 | 0.311 | 2.015 | 0.213 | 2.031 | 0.129 | 2.016 | 0.095 | 2.027 | 0.065 |

**Table TS3. Scenario 2: Carry-over.** Overview of the estimates (of the mean, $\hat{\mu}$, and standard error of the mean, $\hat{\sigma}$) for the different sample sizes for the models: carry-over adjusted parametric model (COAPM), unadjusted Bayesian Network (BN), Bayesian Network with time adjustment (BNt), G-Estimation - independence (GEST-i), G-Estimation - AR1 (GEST-AR1), Linear Model (LM), and Sample Mean (SM).

| **Missing** | **Patients** | 5 | | 10 | | 25 | | 50 | | 100 | |
|---|---|---|---|---|---|---|---|---|---|---|---|
| **Values** | **Method** | $\hat{\mu}$ | $\hat{\sigma}$ | $\hat{\mu}$ | $\hat{\sigma}$ | $\hat{\mu}$ | $\hat{\sigma}$ | $\hat{\mu}$ | $\hat{\sigma}$ | $\hat{\mu}$ | $\hat{\sigma}$ |
| **Without** | **COAPM** | 1.952 | 0.294 | 1.944 | 0.256 | 1.999 | 0.168 | 1.995 | 0.133 | 1.975 | 0.099 |
| | **BN** | 1.633 | 0.428 | 1.542 | 0.305 | 1.602 | 0.187 | 1.607 | 0.133 | 1.588 | 0.083 |
| | **BN-t** | 1.579 | 0.551 | 1.572 | 0.485 | 1.609 | 0.318 | 1.644 | 0.246 | 1.621 | 0.148 |
| | **GEST-i** | 1.500 | 0.244 | 1.535 | 0.234 | 1.592 | 0.126 | 1.604 | 0.098 | 1.581 | 0.071 |
| | **GEST-AR1** | 0.462 | 0.256 | 0.562 | 0.266 | 0.630 | 0.158 | 0.619 | 0.109 | 0.634 | 0.071 |
| | **LM** | 1.585 | 0.287 | 1.533 | 0.231 | 1.592 | 0.126 | 1.604 | 0.098 | 1.581 | 0.071 |
| | **SM** | 1.584 | 0.292 | 1.533 | 0.231 | 1.595 | 0.127 | 1.605 | 0.099 | 1.582 | 0.070 |
| **With** | **COAPM** | 2.150 | 0.510 | 2.159 | 0.346 | 2.180 | 0.228 | 2.174 | 0.178 | 2.198 | 0.104 |
| | **BN** | 1.620 | 0.428 | 1.622 | 0.304 | 1.572 | 0.162 | 1.600 | 0.125 | 1.591 | 0.086 |
| | **BN-t** | 1.633 | 0.535 | 1.650 | 0.436 | 1.623 | 0.380 | 1.601 | 0.231 | 1.634 | 0.165 |
| | **GEST-i** | 1.536 | 0.235 | 1.611 | 0.220 | 1.566 | 0.123 | 1.579 | 0.097 | 1.579 | 0.061 |
| | **GEST-AR1** | 0.822 | 0.377 | 0.810 | 0.313 | 0.767 | 0.170 | 0.791 | 0.115 | 0.788 | 0.079 |
| | **LM** | 1.570 | 0.319 | 1.604 | 0.229 | 1.566 | 0.123 | 1.579 | 0.097 | 1.579 | 0.061 |
| | **SM** | 1.562 | 0.349 | 1.592 | 0.224 | 1.565 | 0.123 | 1.575 | 0.095 | 1.573 | 0.063 |

**Table TS4.** **Scenario 3: Activity interaction.** Overview of the estimates (of the mean, $\hat{\mu}$, and standard error of the mean, $\hat{\sigma}$) for the different sample sizes for the models: carry-over adjusted parametric model (COAPM), unadjusted Bayesian Network (BN), Bayesian Network with time adjustment (BNt), G-Estimation - independence (GEST-i), G-Estimation - AR1 (GEST-AR1), Linear Model (LM), and Sample Mean (SM).

| Missing | Patients | 5 | | 10 | | 25 | | 50 | | 100 | |
|---|---|---|---|---|---|---|---|---|---|---|---|
| **Values** | **Method** | $\hat{\mu}$ | $\hat{\sigma}$ | $\hat{\mu}$ | $\hat{\sigma}$ | $\hat{\mu}$ | $\hat{\sigma}$ | $\hat{\mu}$ | $\hat{\sigma}$ | $\hat{\mu}$ | $\hat{\sigma}$ |
| **Without** | **COAPM** | 1.950 | 0.311 | 1.934 | 0.262 | 1.935 | 0.172 | 1.947 | 0.119 | 1.952 | 0.078 |
| | **BN** | 1.805 | 1.120 | 1.744 | 0.599 | 1.682 | 0.425 | 1.649 | 0.323 | 1.679 | 0.209 |
| | **BN-t** | 1.801 | 1.164 | 1.775 | 0.609 | 1.709 | 0.434 | 1.665 | 0.344 | 1.706 | 0.234 |
| | **GEST-i** | 1.848 | 0.309 | 1.946 | 0.253 | 1.959 | 0.164 | 1.959 | 0.110 | 1.953 | 0.084 |
| | **GEST-AR1** | 1.865 | 0.343 | 1.919 | 0.167 | 1.960 | 0.109 | 1.948 | 0.073 | 1.934 | 0.053 |
| | **LM** | 1.986 | 0.293 | 1.947 | 0.251 | 1.959 | 0.164 | 1.959 | 0.110 | 1.953 | 0.084 |
| | **SM** | 0.203 | 0.130 | 0.163 | 0.122 | 0.091 | 0.066 | 0.067 | 0.048 | 0.047 | 0.036 |
| **With** | **COAPM** | 1.964 | 0.372 | 1.978 | 0.310 | 1.972 | 0.190 | 1.978 | 0.137 | 1.971 | 0.085 |
| | **uBN** | 1.843 | 0.813 | 1.759 | 0.685 | 1.806 | 0.326 | 1.698 | 0.301 | 1.669 | 0.209 |
| | **BNt** | 1.892 | 0.819 | 1.770 | 0.697 | 1.816 | 0.365 | 1.709 | 0.317 | 1.667 | 0.225 |
| | **GEST-i** | 1.974 | 0.231 | 2.008 | 0.265 | 1.975 | 0.158 | 1.981 | 0.115 | 1.972 | 0.072 |
| | **GEST-AR1** | 1.853 | 0.167 | 1.972 | 0.176 | 1.964 | 0.102 | 1.968 | 0.082 | 1.961 | 0.052 |
| | **LM** | 1.962 | 0.342 | 2.000 | 0.272 | 1.975 | 0.158 | 1.981 | 0.115 | 1.972 | 0.072 |
| | **SM** | 0.228 | 0.164 | 0.164 | 0.138 | 0.113 | 0.089 | 0.080 | 0.061 | 0.051 | 0.037 |

**Table TS5.** **Scenario 4: Activity interaction and Carry-Over.** Overview of the estimates (of the mean, $\hat{\mu}$, and standard error of the mean, $\hat{\sigma}$) for the different sample sizes for the models: carry-over adjusted parametric model (COAPM), unadjusted Bayesian Network (BN), Bayesian Network with time adjustment (BNt), G-Estimation - independence (GEST-i), G-Estimation - AR1 (GEST-AR1), Linear Model (LM), and Sample Mean (SM).

| Missing | Patients | 5 | | 10 | | 25 | | 50 | | 100 | |
|---|---|---|---|---|---|---|---|---|---|---|---|
| **Values** | **Method** | $\hat{\mu}$ | $\hat{\sigma}$ | $\hat{\mu}$ | $\hat{\sigma}$ | $\hat{\mu}$ | $\hat{\sigma}$ | $\hat{\mu}$ | $\hat{\sigma}$ | $\hat{\mu}$ | $\hat{\sigma}$ |
| **Without** | **COAPM** | 2.019 | 0.373 | 2.004 | 0.267 | 2.067 | 0.188 | 2.023 | 0.148 | 2.009 | 0.110 |
| | **BN** | 1.701 | 0.847 | 1.409 | 0.555 | 1.364 | 0.427 | 1.365 | 0.349 | 1.366 | 0.256 |
| | **BN-t** | 1.791 | 0.855 | 1.474 | 0.569 | 1.447 | 0.437 | 1.448 | 0.350 | 1.442 | 0.258 |
| | **GEST-i** | 1.885 | 0.648 | 1.641 | 0.256 | 1.660 | 0.177 | 1.636 | 0.120 | 1.637 | 0.091 |
| | **GEST-AR1** | 0.809 | 0.578 | 0.611 | 0.303 | 0.584 | 0.175 | 0.595 | 0.140 | 0.556 | 0.096 |
| | **LM** | 1.636 | 0.391 | 1.641 | 0.255 | 1.660 | 0.177 | 1.636 | 0.120 | 1.637 | 0.091 |
| | **SM** | 0.410 | 0.229 | 0.345 | 0.181 | 0.301 | 0.123 | 0.330 | 0.085 | 0.325 | 0.059 |
| **With** | **COAPM** | 2.212 | 0.563 | 2.213 | 0.443 | 2.234 | 0.305 | 2.229 | 0.223 | 2.169 | 0.207 |
| | **BN** | 1.535 | 0.790 | 1.421 | 0.575 | 1.341 | 0.500 | 1.412 | 0.297 | 1.371 | 0.209 |
| | **BN-t** | 1.596 | 0.804 | 1.507 | 0.590 | 1.422 | 0.502 | 1.488 | 0.292 | 1.452 | 0.207 |
| | **GEST-i** | 1.477 | 0.472 | 1.633 | 0.289 | 1.640 | 0.196 | 1.626 | 0.134 | 1.606 | 0.092 |
| | **GEST-AR1** | 0.644 | 0.353 | 0.742 | 0.324 | 0.690 | 0.178 | 0.710 | 0.140 | 0.720 | 0.100 |
| | **LM** | 1.649 | 0.395 | 1.636 | 0.290 | 1.640 | 0.196 | 1.626 | 0.134 | 1.606 | 0.092 |
| | **SM** | 0.388 | 0.265 | 0.375 | 0.222 | 0.312 | 0.153 | 0.329 | 0.101 | 0.342 | 0.072 |
